# COVID-19 vaccine incentive scheduling using an optimally controlled reinforcement learning model

**DOI:** 10.1101/2022.02.17.22271145

**Authors:** K. Stuckey, P.K. Newton

## Abstract

We model Covid-19 vaccine uptake as a reinforcement learning dynamic between two populations: the vaccine adopters, and the vaccine hesitant. Using data available from the Center for Disease Control (CDC), we calculate a payoff matrix governing the dynamic interaction between these two groups and show they are playing a Hawk-Dove evolutionary game with an internal evolutionarily stable Nash equilibrium (the asymptotic percentage of vaccinated in the population). We then ask whether vaccine adoption can be improved by implementing dynamic incentive schedules that reward/punish the vaccine hesitant, and if so, what schedules are optimal and how effective are they likely to be? When is the optimal time to start an incentive program, and how large should the incentives be? By using a tailored replicator dynamic reinforcement learning model together with optimal control theory, we show that well designed and timed incentive programs can improve vaccine uptake by shifting the Nash equilibrium upward in large populations, but only so much, and incentive sizes above a certain threshold show diminishing returns.

## I. INTRODUCTION

The voluntary uptake of vaccines for Covid-19 has proven to be a challenge across the world, but particularly in the United States where multiple vaccine options have been available since early in 2021. After an initial population of early adopters were vaccinated, the surge began to slow, despite widespread availability, and has now reached what looks to be a fairly stable resting point (see figure 1). While 100% voluntary compliance is rarely if ever achievable, vaccine hesitancy [1] has proven to be more widespread for Covid-19 than for other vaccines, such as seasonal flu vaccines [2], the polio vaccine, smallpox, HPV and others [3]. With any widespread nationally coordinated vaccination effort, there will always be a population of people who we label vaccine adopters (e.g. elderly, immuno-compromised, healthcare workers) who get vaccinated as soon as they are eligible, or shortly thereafter, then others follow. The vaccine uptake curve for this group in these early stages is limited mostly by vaccine availability and logistics. There is also a vaccine hesitant population who will delay their initial chances to get vaccinated, then as they see others getting sick and weigh evidence and public opinion, some might decide to vaccinate (adopters), while still others might further delay, or forgo their chance altogether for various reasons (hesitant) [4]. We view the full population as a collection of two types of players in a time-evolving game, who interact, learn, and receive payoffs (reward/punishment) according to the strategy (adopt/forgo) they choose, where the interactions determine the fitness of the players using one of two strategies, and the survival of that strategy is determined by the fitness function. The two competing behaviors ultimately result in a growth curve describing vaccine uptake, shown in figure 1, that starts out rapidly (exponential), then slows down, passing through an inflection point to a fairly stable resting percentage of vaccinated individuals, which in the United States seems to have settled at just under 60% of the population (figure 1).

**FIG. 1.**
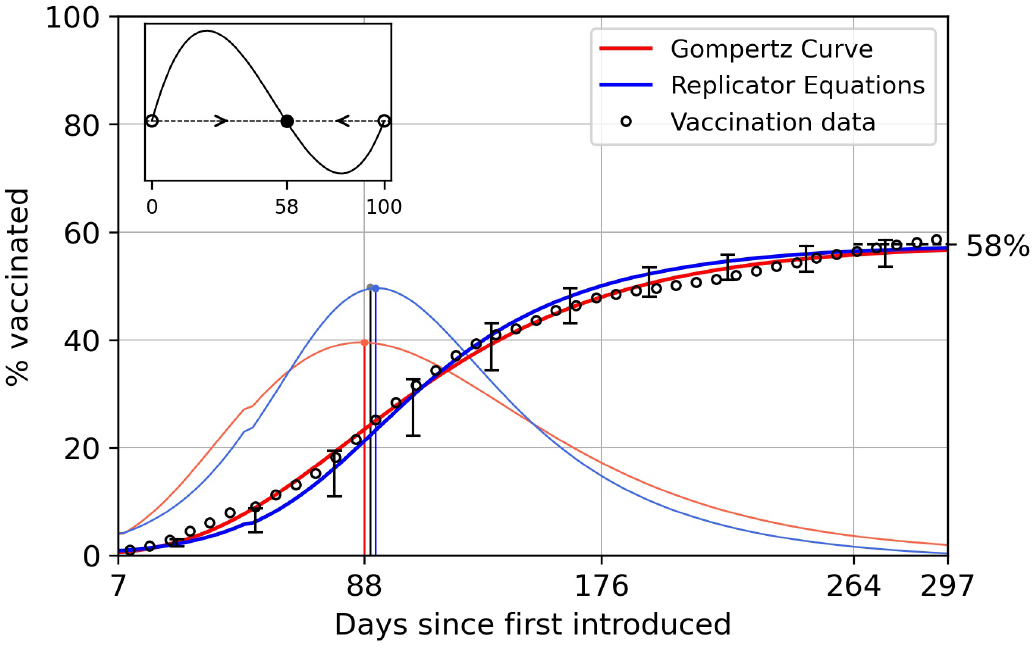
Covid-19 vaccine uptake data in the United States (1 dose for Johnson & Johnson or 2 doses for Pfizer/Moderna) starting in January 2021. Red curve shows a Gompertzian fit to the data (first derivative curve also in red), blue curve shows the results of the Hawk-Dove evolutionary game theory model (first derivative curve also in blue). Error bars show one standard deviation from the mean using an ensemble of stochastic realizations from the model. Vertical lines mark the inflection point (maxima of the first derivative) where vaccine uptake begins to slow, resulting in an asymptote (Nash equilibrium) at roughly 58% of the population. Upper left insert shows the phase-plane diagram for the Hawk-Dove dynamical system with an internal evolutionary stable state (ESS) at 58%. How much can well designed incentive programs push this percentage up?

The question we address in this paper is whether or not a well designed (optimized) punishment/reward system can significantly alter this natural dynamic, and if so, how best to do that? Vaccine incentive programs have been utilized with varying degrees of success for other vaccines, but for the Covid-19 vaccine they have largely been local (county-wide and state-by-state) and somewhat haphazard, ranging from small cash rewards handed out at vaccination clinics, medium-sized vacation add-ons, or larger lottery-style rewards [5, 6]. Punishments for the unvaccinated have also been levied, ranging from the small extra hassle of requiring weekly Covid testing, more severe restrictions of not being allowed entry to restaurants or public events, and larger vaccine mandates that require vaccines as part of the employment requirement or school enrollment [7–9] Table 1 shows a compilation of the mostly ad hoc strategies that have been implemented in states across the country but larger scale national programs have not been systematically designed or implemented. Specific questions we address in this paper include whether there are inherent limitations to well designed (optimized) punishment/reward systems, if implemented on a wide-scale basis? Are there optimal schedules that can be designed that would work most effectively? What are the optimal starting and ending times for such dynamic incentive programs? Is there a point of diminishing returns where larger incentives are no longer as effective? By modeling the vaccine uptake problem as a reinforcement learning evolutionary game played between two sub-populations of “players” (the vaccine adopters and the vaccine hesitant), we address these questions within the context of a mathematical model calibrated with vaccine uptake data obtained from the Center for Disease Control both on a nationwide level, and a state-by-state level. With models tailored to individual states and for different age groups, we are able to test various types of incentive schedules to produce upper and lower bounds (using the Pontryagin maximum/minimum principle from optimal control theory) on the inherent limitations of dynamic incentive programs, and by producing incentive/response curves (analogous to chemotherapeutic dose/response curves [10]), we are able to hypothesize likely responses to different types and sizes of the incentive schedules.

**TABLE I.**
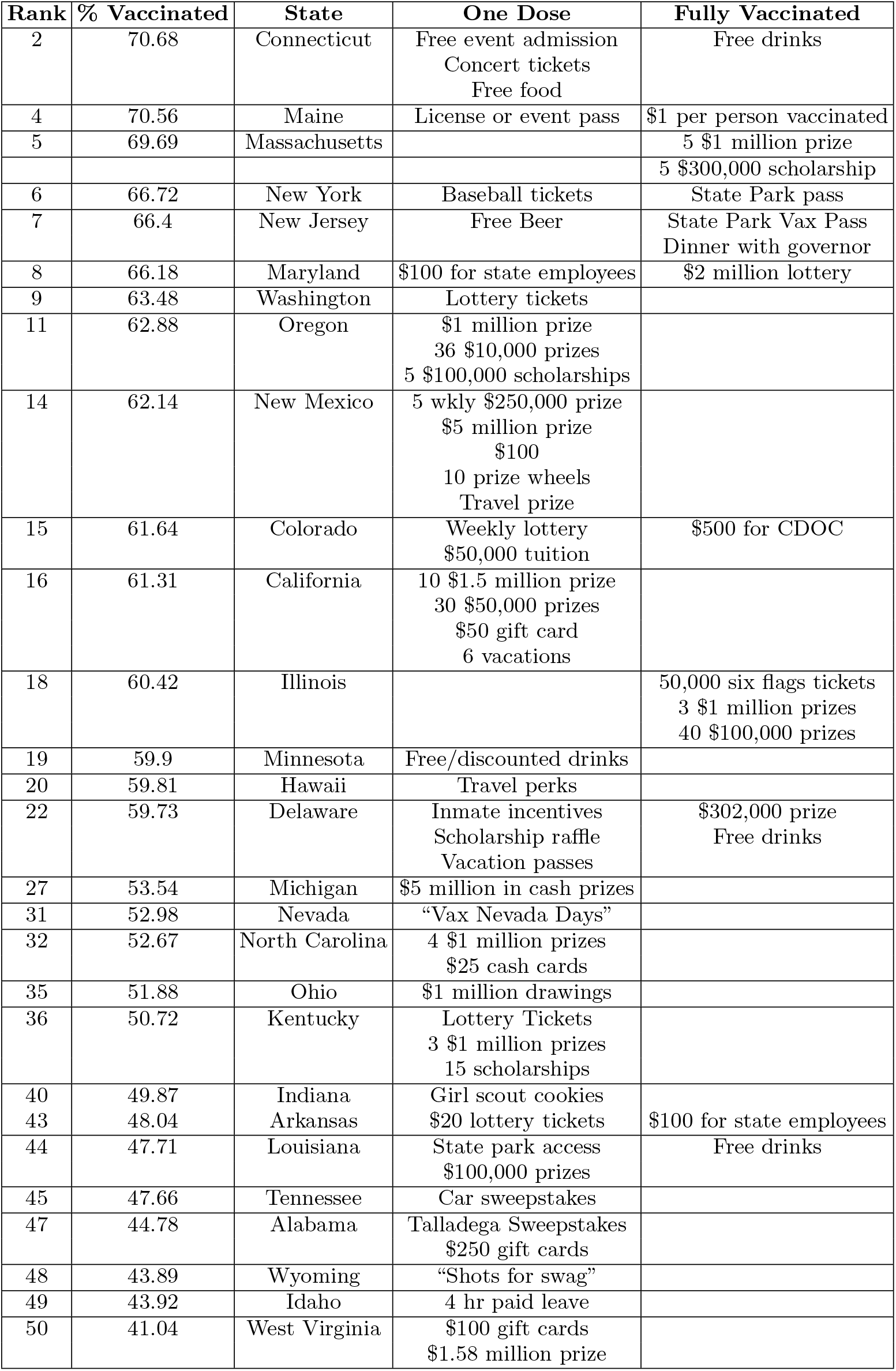
Compiled information on different forms of vaccine incentives states have used. States are ranked in order of highest adoption percentage to lowest. Not listed are states where we were not able to obtain information on any incentive programs.

Aspects of vaccine policy and individual decision making surrounding these policies has been studied recently using mathematical models. Korn et al. [11] has studied vaccine uptake arguing that it can be viewed as a social contract where individuals reward others who comply and punish those who do not, while Bauch et al. [12] frame it similarly in terms of the complex trade-offs between group interests versus self-interest arguing that, in the case of extreme events (e.g. bio-terrorist attack), it is unlikely that voluntary vaccination levels alone would reach the group optimal level necessary for herd immunity. Bauch et al. [13, 14] have used game-theoretical models to help explain human decision-making surrounding vaccine uptake studying how vaccine scares unfold [15], and in [16], they invoke imitation dynamics models to understand the complex interplay between vaccine coverage, disease prevalence, and individual decision making. More general modeling frameworks have used tools borrowed from statistical physics in interesting ways [17] to model vaccine dynamics.

Our approach makes use of the vaccine uptake data (country-wide data, and four key states) available at https://covid.cdc.gov/covid-data-tracker/#datatracker-home to fit the Gompertzian parameters (*a, b, c*) and the entries of the 2 × 2 payoff matrix that describes the evolutionary game played between the vaccine adopters and the vaccine hesitant populations. The data shows that the population is effectively playing a Hawk-Dove game with an evolutionary stable internal fixed point (ESS) representing the percentage of vaccine adopters (Hawks) in the population. We then use optimal control theory to design time-dependent incentive schedules that alter the baseline payoff matrix entries (altering the reward/punishment balance) in order to obtain upper (and lower) bounds on how different incentive strategies can shift the asymptotic percentage of vaccine adopters in the population. This control technique was originally developed for the design of adaptive/optimal chemotherapy schedules for controlling resistance in tumors [18–23]. Here, we exploit the observation that optimizing vaccine incentive schedules is analogous to optimizing chemotherapy schedules to produce dose-response curves [10] for specific goals, such as, for example, avoiding chemotherapeutic resistance [18–21]. The adoption of these techniques to vaccine incentive scheduling presents a different set of questions and challenges but can be addressed within a similar modeling framework. Other recent work that makes use of feedback control ideas to develop COVID-19 policies includes [24]. While the merging of reinforcement learning models with optimal/adaptive control theory is a new and promising field with many potential applications, a nice introduction to the field, described mostly in the robotics framework, can be found in a recent monograph [25].

## II. THE VACCINE UPTAKE MODEL

### Calibrating the Gompertzian curves

The vaccine uptake curve shown in figure 1 is a three parameter (*a, b, c*) Gompertzian curve,

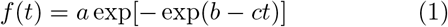

which has a long history of use in actuarial sciences (laws of human mortality), economics (growth laws of wealth), biology (population growth and saturation), cancer (tumor growth) [26, 27]. Key parameters for fitting such a curve to this data are: (1) *T* : the location of the inflection point (shown in figure 1); (2) *f*′(*T*): the slope of the tangent line at the inflection point (the growth rate when the growth curve changes from concave up to concave down); (3) *f*′(0): the slope of the tangent line at the origin (initial growth rate). In eqn (1), *a* is the asymptote (lim_*t*→∞_*f*(*t*)) (also known as carrying-capacity in other contexts [28, 29]), *b* is the displacement along the *t*-axis (time-shift parameter), and *c* is a time-scaling factor. In terms of those parameters, the inflection point is located at *T*, where:

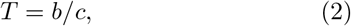

is the ratio of time-shift to time scaling parameters, while the slope of *f*(*t*) at the origin and inflection points are given by:

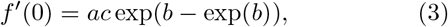

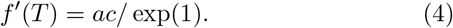

We fit the three parameters (*a, b, c*) to the data as shown in figure 1 (in red) for the US population. For this, the asymptote is roughly 58% vaccinated, and the uptake inflection point is *T* ≈ 88 days from when vaccines first became available. Table 2 provides a summary of all of the model parameters we use for the different subpopulations. With these parameters, we develop the reinforcement learning model.

**TABLE II.** Model parameters for different population groups.

| Population | $T$ (days) | $f'(0)$ | $f'(T)$ |
| --- | --- | --- | --- |
| US population | 88 | 0.028256504 | 0.405499161 |
| 18 – 39 year olds | 106 | 0.008343649 | 0.382741771 |
| 40 – 64 year olds | 90 | 0.004432806 | 0.597362637 |
| 65+ year olds | 58 | 0.015256548 | 1.024397092 |
| Connecticut | 86 | 0.012707894 | 0.563297 |
| Vermont | 89 | 0.002882054 | 0.612739997 |
| Idaho | 77 | 0.030170215 | 0.327559854 |
| West Virginia | 63 | 0.065387774 | 0.331017921 |

### The reinforcement learning/replicator model

We use the replicator dynamics equations from evolutionary game theory and reinforcement learning to model vaccine uptake dynamics between the two populations *x*_*A*_ (vaccine adopters) and *x*_*H*_ (vaccine hesitant), where each represents a proportion of the entire population, 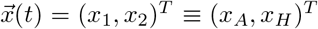 ; *x*_*A*_ + *x*_*H*_ = 1. The essential feature of replicator dynamics is that people (reinforcement learners) copy others, and succesful strategies get replicated more frequently than unsucessful strategies [30]. This makes the model useful not only in contexts where Darwinian evolution by natural selection is prominant [31], as cells and organisms with higher fitness (measured by their ability to replicate) more often pass along their genetic characteristics in the population, but also in any reinforcement learning setting where learners copy succesful strategies [32] more often than unsuccesful ones (success begets success and failure spirals downward), with success determined by fitness level. The attractiveness of this framework in the present context is that it has been widely documented that vaccine uptake is more common in a positive uptake environment, and less common in settings where fewer people choose to get vaccinated. This dynamic is the hallmark of a reinforcement learning process where people interact, learn strategies from others, receive payoffs (in the form of advantage or disadvantage) based on strategies they adopt, the payoffs determine the fitness (ability to survive) of those strategies in the overall population, the fitness controls the survival probability of the strategy.

To formulate the dynamical system, we use:

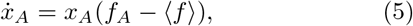

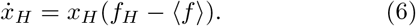

Here, *f*_*A*_ and *f*_*H*_ denote the fitness of the vaccine adopters and the vaccine hesitant populations, while ⟨*f*⟩ denotes the average fitness of the entire population under consideration. The system simply says that the growth rate of each sub-population 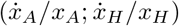 is governed by the difference between the fitness of that population and the overall average fitness of both populations. The more each sub-population fitness deviates from the average (either above or below), the larger/smaller the instantaneous growth rate is of that strategy in the population. The fitnesses are defined via a 2 × 2 payoff matrix *A* as:

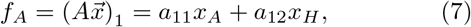

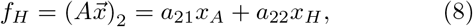

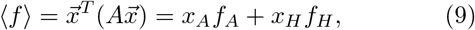

with:

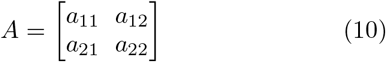

which defines the evolutionary game being played as determined by the CDC data. *The four entries of this matrix encode the punishment-reward balance (i*.*e. payoffs) associated with competition between the two groups and is the heart of the model*. As described in [33], the payoffs are decided by many complex factors, including each person’s perceived risk of infection (which can vary in time), the severity of the disease (which can vary in time), perhaps measured in hospitalization rates, financial costs of vaccinations, and also the perceived uptake of vaccinations by others. Increasing/decreasing either of the entries of the top row of *A* increases/decreases the fitness of the vaccine adopter population, whereas increasing/decreasing either of the entries of the bottom row of *A* increases/decreases the fitness of the hesitant population. Without loss, we can choose *a*_22_ = 0, while the remaining three entries can be obtained as functions of (*a, b, c*) which were optimally fit to the data. Thus, all of the complexities associated with the many decision processes which result in the Gompertzian uptake curves are neatly packaged into three of the four entries of the payoff matrix determining the evolutionary game which unfolds between the vaccine adopters and the vaccine hesitant. This gives rise to a Hawk-Dove evolutionary game based on the inequalities: *a*_21_ *> a*_11_ *> a*_12_ *> a*_22_ = 0. A key feature of a Hawk-Dove evolutionary game is the existence of an internal ∈ (0, 1) ESS (Nash equilibrium), which we denote by *a* (the asymptote of *f*(*t*)). As shown in figure 1, for the US population as a whole, *a* ≈ 0.58. Figure 2 shows the data, curve fit, and replicator dynamic model for the two states with the highest vaccine uptake percentages (Vermont and Connecticut ≈ 69%), and the two lowest (West Virginia and Idaho ≈ 41%). Figure 2(a) shows the results of the vaccine uptake data along with both the Gompertzian curve fit and the replicator dynamics model for the four states, along with the entire US population. In figure 2(b) we break the US data into three different age groups (18-39; 40-64; 65+) in a similar way that the vaccine rollout prioritized these groups. This is reflected in the leftward shift of the curve corresponding to the older compared to younger groups, with the oldest population showing the steepest uptake curve consistent with the notion that this group was among the most eager to be vaccinated. The inflection point for the US population as a whole is roughly at *T* ≈ 88 days which we take as a benchmark for scaling time when we implement our control strategy on this group. Similarly, for all other subgroups, we use the corresponding inflection point location associated with that subgroup (see Table 2).

**FIG. 2.**
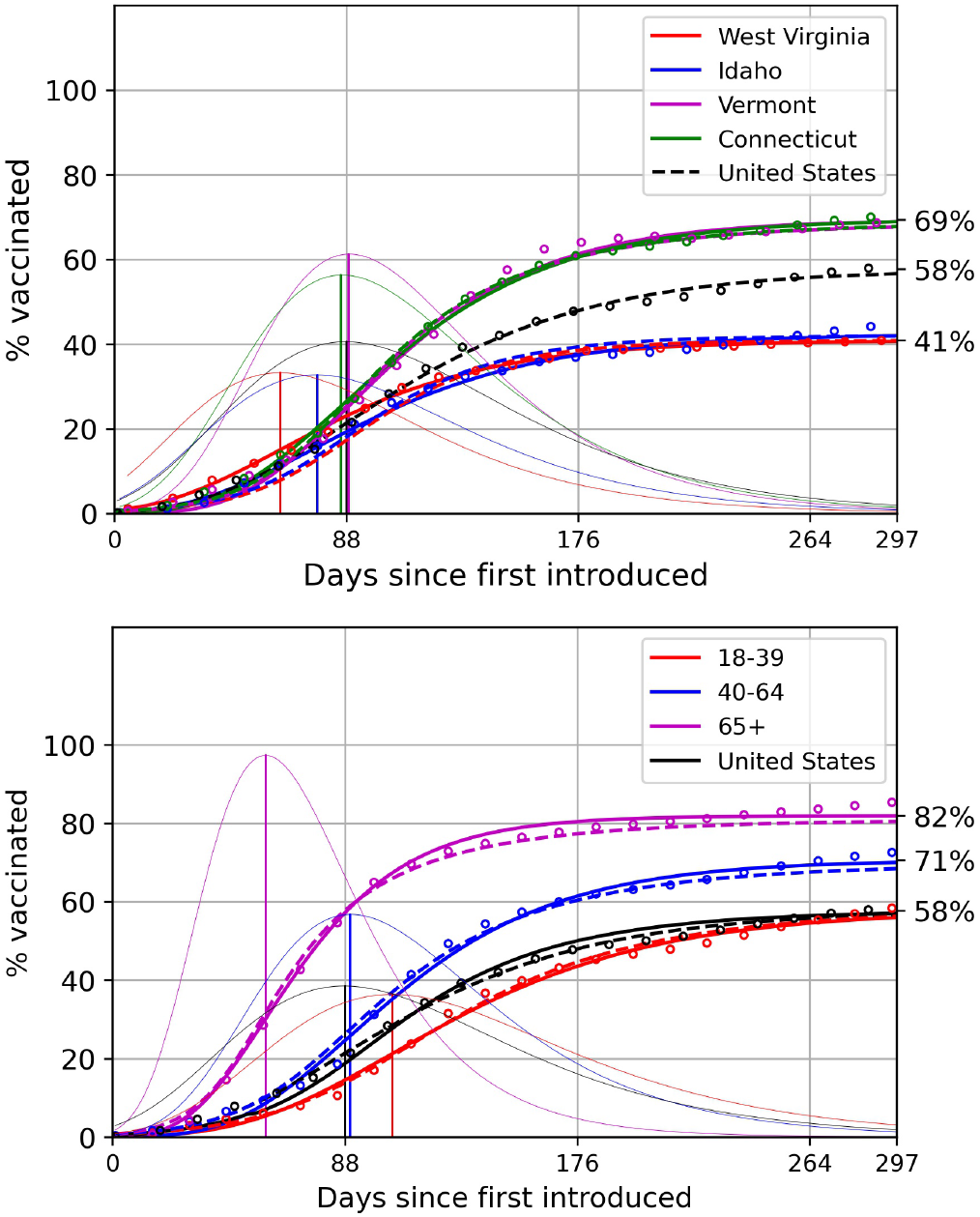
Vaccine uptake curves (data (dots), Gompertzian fit (dashed), replicator model (solid)) of different groups within the United States. Vertical lines mark inflection points at maximizers of the derivative curves. (a) The two states with highest vaccination rate, Vermont and Connecticut, versus two states with lowest vaccination rates, Idaho and West Virginia; (b) Three different age groups: 18-39, 40-64 and 65+;

### Shifting vaccine uptake curves with time-dependent payoffs

To implement an optimal vaccine incentive strategy, we now consider the time-dependent payoff matrix:

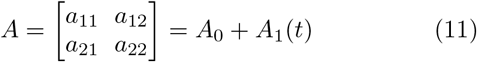

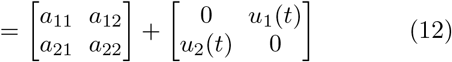

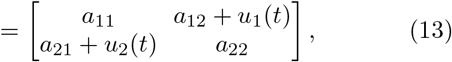

where *A*_1_(*t*) represents our control with entries in the off-diagonal terms (without loss of generality), and *A*_0_ is the baseline Hawk-Dove payoff matrix as obtained from the vaccine uptake data. The time-dependent controllers 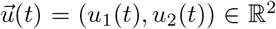 are bounded above and below (based on the incentive size *p*):

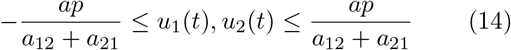

(0 ≤*p* ≤ 1) and a global contraint on the incentive schedule, 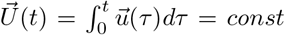. is enforced, all of which play an important role in determining the optimal incentives.

## III. RESULTS

### State-by-state results

The first question we address using our optimized incentive model, is whether it is possible to incentivize the states with low vaccine uptake curves (West Virginia and Idaho) to bring them up to the level of states with high uptake curves (Vermont and Connecticut). Figure 3(a) shows the result of our simulations for West Virginia. With relatively large incentize sizes roughly between 15% − 20% (measure normalized by the baseline value), we show this is possible. But we consider this range of incentive sizes to be so large that the price of implementing them might be prohibitive. Figure 3(b) shows our simulations for Idaho, with the same general conclusions as West Virginia. Incentive sizes this large can have a considerable effect, but the price would be high to implement them.

**FIG. 3.**
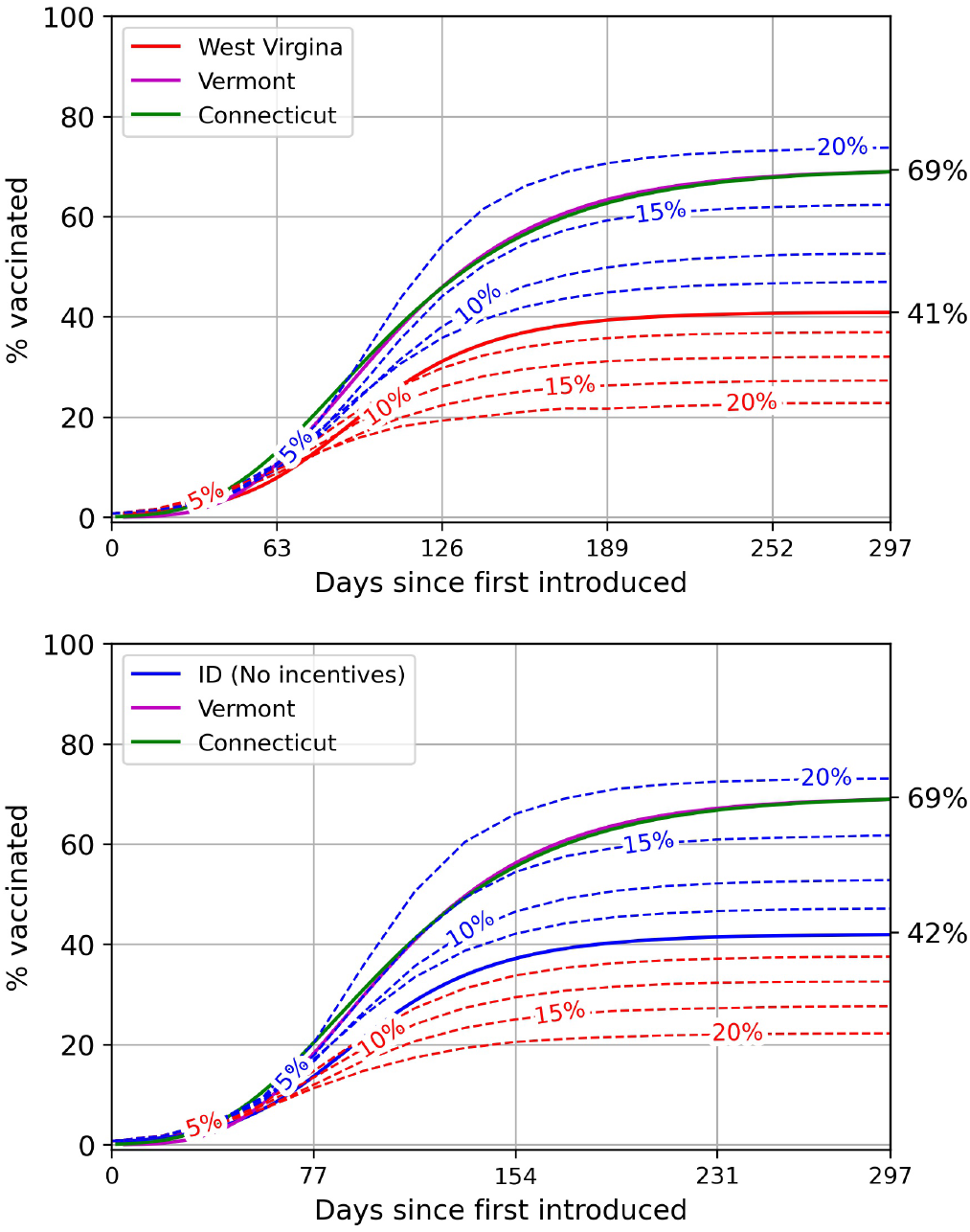
Maximizing (blue) and minimizing (red) vaccination percentages for state with lowest vaccination rates, West Virginia and Idaho, in comparison with states with the highest vaccination rates, Vermont (purple) and Connecticut (green). (a) Solid red curve depicts West Virginia vaccine uptake curve. Blue dashed curves show West Virginia model with optimized incentive schedules (upper bound) using 5%, 10%, 15%, 20% incentive sizes (normalized uisng baseline value). Red dashed curves show West Virginia model with optimized incentive schedules (lower bound) using 5%, 10%, 15%, 20% incentive sizes (normalized uisng baseline value);(b) Solid blue curve depicts Idaho’s vaccine uptake curve. Blue dashed curves show Idaho model with optimized incentive schedules (upper bound) using 5%, 10%, 15%, 20% incentive sizes (normalized uisng baseline value). Red dashed curves show Idaho model with optimized incentive schedules (lower bound) using 5%, 10%, 15%, 20% incentive sizes (normalized using baseline value).

### Optimal timing

We next address whether or not the initial start-time of our optimal incentive schedule has much impact on the end result. The short answer to this is no, it does not, as shown in the figure 4 panel for the (a) US population, (b) Connecticut population, (c) Idaho population, and (d) West Virginia population. In all cases, the incentivized curves (dashed) asymptote to the absolute max/min curves no matter when the schedules begin. This indicates that we could begin the schedules at the inflection point of the uptake curves, allowing us to collect and develop the model in real time as the uptake dynamics unfolds, designing the optimal incentive schedules to use going forward. The one caveat with this approach is that although the curves all reach the same asymptote, if time is of the essence (say because of high death rates in the unvaccinated population), there could well be advantages to starting the incentive schedules as early as possible. To design optimal schedules in real time before reaching the inflection point of the uptake curve would require a seperate careful forecasting model based only on earlier data.

**FIG. 4.**
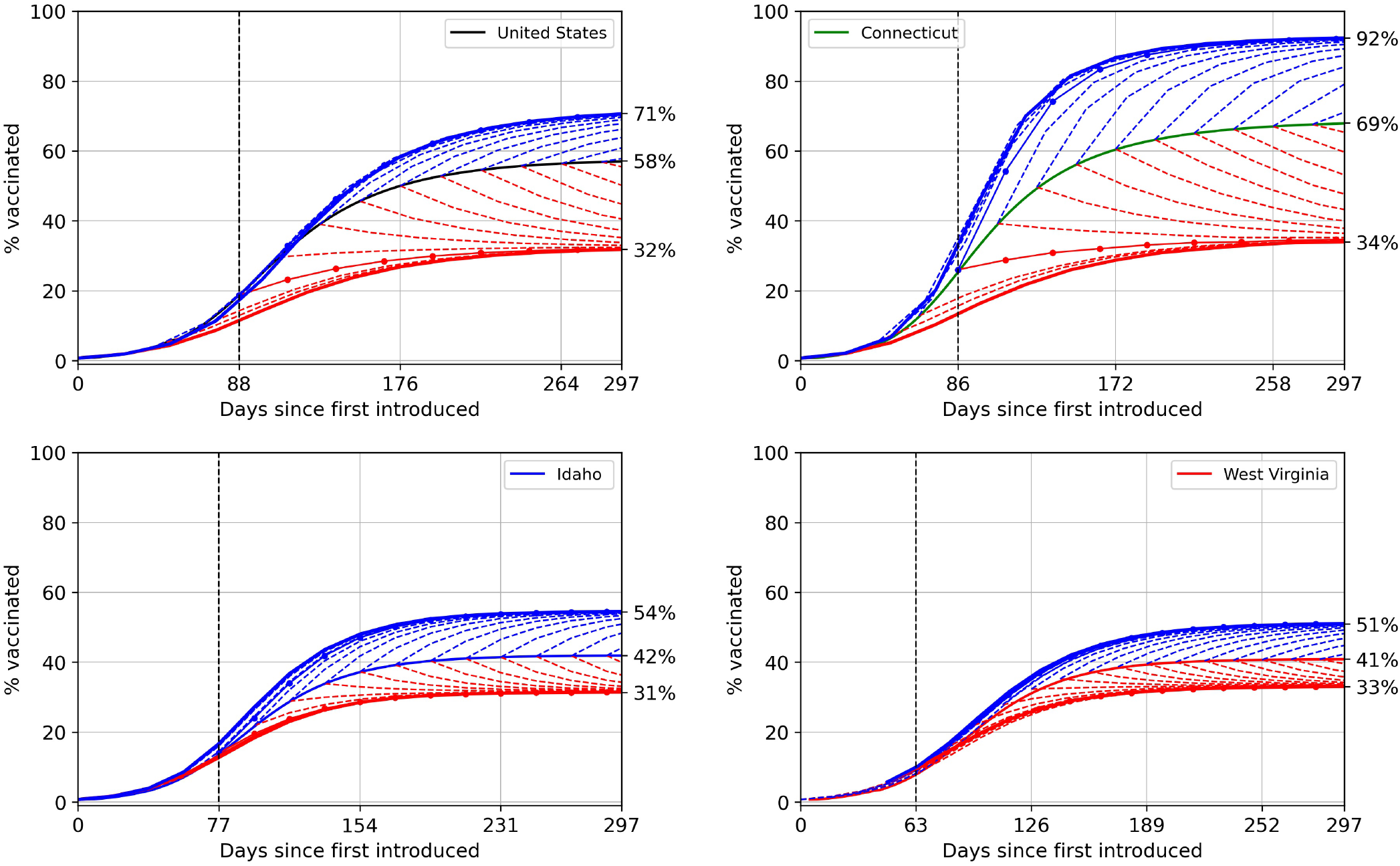
Maximizing and minimizing vaccination uptake with controllers turning on at different times. All plots show incentive shedules beginning at 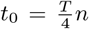 for increasing *n*. In all cases, the incentivized schedules reach upper and lower asymptotes, indicating relative insensitivity of the optimal outcome to start times. (a) United States un-incentivized uptake curve (black), *T* = 88 days. Blue shows optimized upper bounds using incentive schedules, red shows lower bounds using incentive schedules; (b) Connecticut un-incentivized uptake curve (green), *T* = 86 days. Blue shows optimized upper bounds using incentive schedules, red shows lower bounds using incentive schedules; (c) Idaho un-incentivized uptake curve (blue), *T* = 77 days. Blue shows optimized upper bounds using incentive schedules, red shows lower bounds using incentive schedules; (d) West Virginia un-incentivized uptake curve (red), *T* = 63 days. Blue shows optimized upper bounds using incentive schedules, red shows lower bounds using incentive schedules.

### Incentive-response curves

We now address the question of what incentive size leads to the best response? Figure 5 shows the percent shift in the asymptote (US population) for different size incentives, both upper (blue) and lower (red) bounds. With no incentive (0%), the asymptote remains at 58% of the population, as expected. In general, the larger the incentive, the larger the response, but two features stand out from these curves. First, low percentage incentives, generally, are more efficient than higher percentage incentives, whose curves all bunch together near an asymptote that even the lower percent incentives generally eventually will reach. The high percentage incentives do have much faster initial rise, so as stated earlier (for different reasons), if time to maximum effect is of the essence, then higher percentage incentives will be superior.

**FIG. 5.**
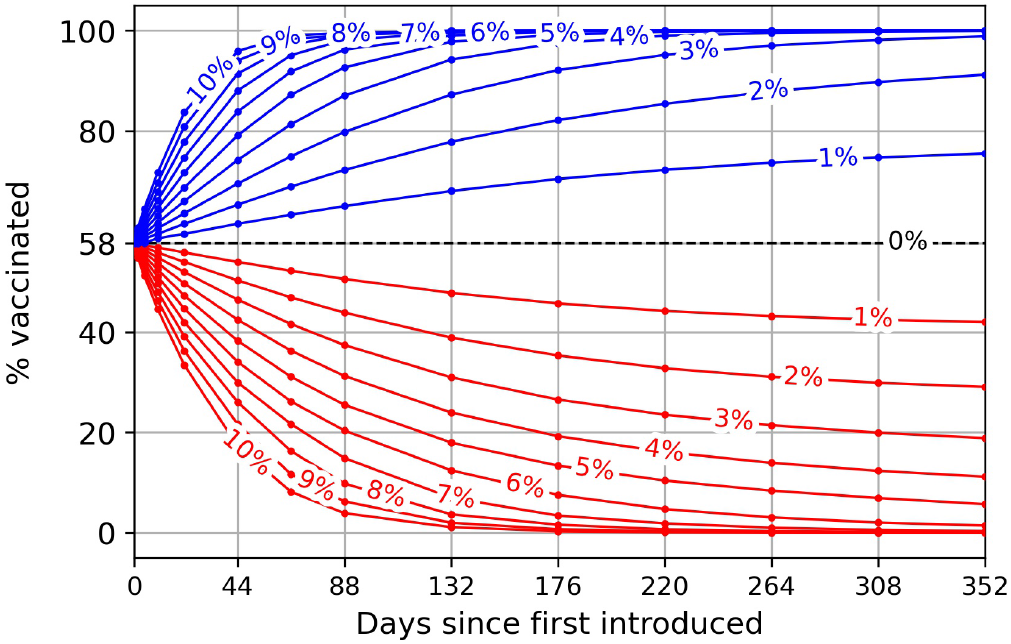
Absolute maximizers (blue) and minimizers (red) for a range of incentive intensities using US population model. Cycle times 44 days *≤ T ≤* 352 days.

### Diminishing returns

Is there a point of diminishing returns on implementing larger incentives, after which the response diminishes? Figure 6 shows an incentive-response curve for the US population. Our model produces a curve (data points fit to three-parameter Gompertzian curve) depicting the incentive strength (abscissa) versus the change in asymptote (ordinate). For incentive strengths below 11%, the curve is concave up, indicating a better response with higher incentives. Above 11%, however, the curve is concave down, indicating a weaker response to higher incentives. We can think of this threshold value (∼11%) as a point of diminishing returns. This is in many ways analogous to dose-response curves in chemotherapy settings [10] where at low values, higher doses produce better results, but past a threshold (typically called maximum tolerated dose, determined via clinical trials) increasing the dose further shows a diminished response. This leads to a theshold value of optimal incentive size, which our model predicts is roughly 11%. We place more value in showing that such a threshold exists in our model, than the actual threshold value, which can be tricky to pin down accurately and actually implement.

**FIG. 6.**
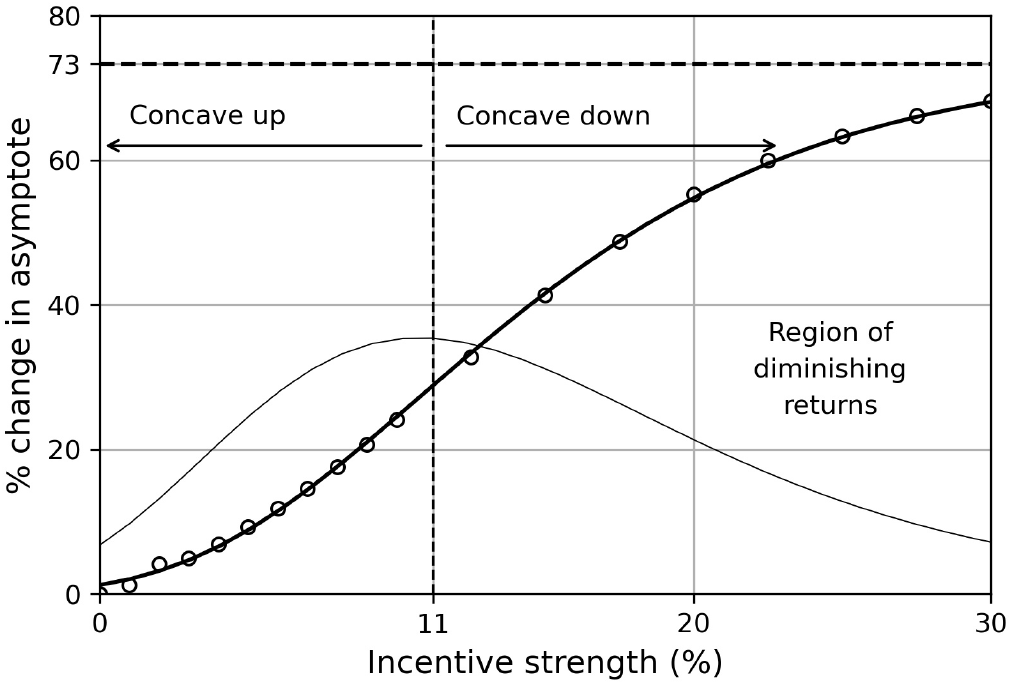
Incentive-response curve fit to data. Abscissa indicates the incentive size (measured as % of baseline). Ordinate shows the shift in the asymptote (measured as % of baseline). Also shown is the first derivative of the response curve. Response curve is concave up for incentive strengths below 11%, and concave down for larger incentives indicating diminishing returns in terms of response.

## IV. DISCUSSION

Although it is presumably unrealistic to assume that optimality will actually be achievable in practice, optimal control nonetheless gives clear upper and lower bounds on what is theoretically attainable in an ideal setting. But there are several tangible ways the model could be improved. First, we make the simplifying assumption that response times to incentives are instantaneous. Building in finite-time responses (i.e. time delays) would make the model more realistic. Second, the hesitant population could be further sub-divided into groups, such as hesitant but willing, hesitant and unwilling, with incentives influencing each of those groups differently. This would lead to a higher dimensional model with more complexity but perhaps higher fidelity. Third, the model assumes what is called a well-mixed population (i.e. no spatial structure). A spatially dependent model would be significantly more complex but has the potential to be more targeted geographically. Finally, the psychological aspects of how people, states, groups, respond to different incentives is not considered in our model. Matching the size of our controllers with actual incentives/punishments would best be handled by experts in human psychology and is not addressed in our approach.

Two strengths of our model we would like to emphasize are that only data up until the inflection point needs to be used, and starting the incentives after that point will ultimately lead to the same shift in the asymptotic percentage of vaccinated people as would have happened if the incentives started earlier. It is not a priori clear whether or not nationwide, state-wide, or even more localized data is most useful, but models that use more localized information (at least state-wide) would probably be more useful as it seems probable that different regions of the country would respond differently to different kinds of incentives.

## V. CONCLUSION

Every vaccine rollout associated with each new epidemic will have its own natural uptake curve, depending esentially on the complex nature of the interactions between the vaccine adopters and the vaccine hesitant populations, and also the interactions within each group, all of which are nicely encoded as elements of the payoff matrix as determined by the data. But it is not unreasonable to speculate that they should all commonly follow the general form of a three-parameter Gompertzian, with different parameters in each case, and geographic location (targeted population), but of the same universal form. This general form is an outcome of the fact that there are, generally speaking, early adopters, followed by a population of players who decide to adopt as time proceeds, leaving only the most hesitant who remain towards the later stages of a rollout. As a vaccine rollout unfolds, the key parameters to obtain from the vaccine uptake curve are: (i) the initial rate of uptake (which we write as % of the relevant population per day), (ii) the inflection point location on the uptake curve (i.e. when uptake begins to slow down), and (iii) the slope of the tangent line at the inflection point (rate of uptake). As long as reliable data is available up until the inflection point, the reinforcement learning model described can then be developed and calibrated in real-time. When the uptake rate begins to slow (i.e. at or near the inflection point), using the controlled replicator dynamical system model, vaccine incentive schedules can be optimized going forward and likely responses can be predicted from the dose-response curves produced by the model. A recent review paper [33] has highlighted the importance and need for using game theory and mathematical models in designing vaccine policy, which we enthusiastically endorse and feel is an under-utilized tool in the arsenal of developing science-based decision making during an infectious outbreak. The framework developed here, which allows the mathematical models to be tailored to specific settings, offers the possibility of testing different strategies in real time for many different scenarios and is flexible, generalizable, relatively simple, and actionable.

## VI. MATERIALS AND METHODS

To implement the Pontryagin maximum (minimum) principle with boundary value constraints in order to compute upper (maximum principle) and lower (minimum principle) bounds, we follow [34] and use:

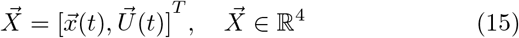

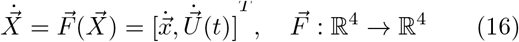

where we would like to minimize or maximize a general cost function:

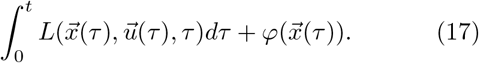

Since we are optimizing the final value, 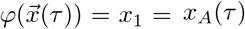 only (i.e. the asymptotic vaccine acceptance value), we can take *L* = 0 (called a Meyer problem [34] developed in the context of missile guidance problems where final distance from the target is minimized). We briefly describe the basic framework and refer readers to [34] for more details on how to implement the approach. In particular, we construct the control theory Hamiltonian:

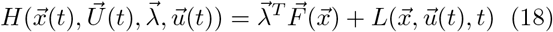

where 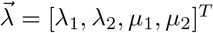 are the co-state functions (i.e. momenta) associated with 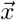 and 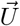 respectively. Assuming that 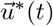 is the optimal control for this problem, with corresponding trajectory 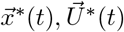, the canonical equations satisfy:

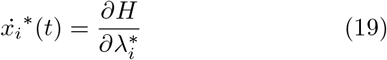

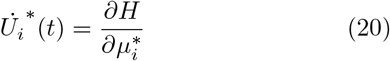

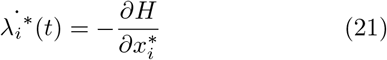

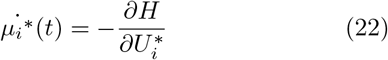

where *i* = (1, 2). The corresponding boundary conditions are:

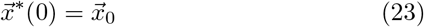

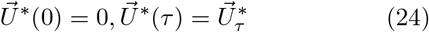

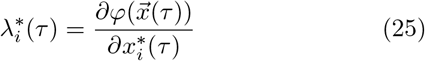

Then, at any point in time, the optimal control 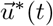 will minimize the control theory Hamiltonian:

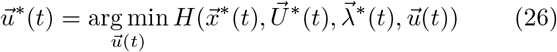

The optimization problem becomes a two-point boundary value problem (using (23)-(25)) with unknowns 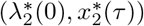 whose solution gives rise to the optimal trajectory 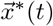 (from (19)) and the corresponding control 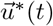 that produces it.

## Data Availability

All data produced are available online at: https://covid.cdc.gov/covid-data-tracker/#datatracker-home

## ACKNOWLEDGMENTS

We gratefully acknowledge support from the Army Research Office MURI Award #W911NF1910269 (2019-2024).

